# PodGPT: An audio-augmented large language model for research and education

**DOI:** 10.1101/2024.07.11.24310304

**Authors:** Shuyue Jia, Subhrangshu Bit, Edward Searls, Meagan V. Lauber, Lindsey A. Claus, Pengrui Fan, Varuna H. Jasodanand, Divya Veerapaneni, William M. Wang, Rhoda Au, Vijaya B. Kolachalama

**Author notes:** These authors contributed equally to this work (Shuyue Jia, Subhrangshu Bit, Edward Searls). Corresponding author: Vijaya B. Kolachalama, PhD.

## Abstract

The proliferation of scientific podcasts has generated an extensive repository of audio content, rich in specialized terminology, diverse topics, and expert dialogues. Here, we introduce a computational framework designed to enhance large language models (LLMs) by leveraging this informational content from publicly accessible podcast data across science, technology, engineering, mathematics and medical (STEMM) disciplines. This dataset, comprising over 3, 700 hours of audio content, was transcribed to generate over 42 million text tokens. Our model, PodGPT, integrates this wealth of complex dialogue found in audio podcasts to improve understanding of natural language nuances, cultural contexts, as well as scientific and medical knowledge. PodGPT also employs retrieval augmented generation (RAG) on a vector database built from articles in Creative Commons PubMed Central and *The New England Journal of Medicine*, enhancing STEMM research and education by providing real-time access to emerging scientific literature. Evaluated across multiple benchmarks, PodGPT demonstrated an average improvement of 3.51 percentage points over standard open-source benchmarks and 3.81 percentage points when augmented with evidence from the RAG pipeline. Moreover, it showcased an average improvement of 4.06 percentage points in its zero-shot multi-lingual transfer ability, effectively generalizing to different linguistic contexts. By harnessing the untapped potential of podcast content, PodGPT advances natural language processing and conversational AI, offering enhanced capabilities for STEMM research and education.

---

The rise of generative artificial intelligence (AI), particularly large language models (LLMs), has marked a transformative shift in data analysis, interpretation, and content generation. These models, trained on extensive textual datasets, have demonstrated the ability to generate contextually accurate and linguistically rich outputs, with profound implications for fields such as science and medicine, where models like OpenAI’s GPT-4 have shown remarkable aptitude ^1–3^. However, the full potential of LLMs in science, technology, engineering, mathematics, and medicine (STEMM) remains under-explored, particularly in integrating non-traditional data modalities such as audio content. Podcasts, which have proliferated across STEMM disciplines, present an untapped repository of expert knowledge, diverse terminologies, and emerging topics. The conversational nature of these recordings encapsulates domain-specific language and dialogue patterns, providing an opportunity to augment language models with rich, real-time, and contextually refined data. By integrating this dynamic source of information, language models could achieve greater precision and relevance within STEMM fields, enhancing their ability to handle complex topics, interdisciplinary discourse, and evolving knowledge landscapes inherent to STEMM.

Recent advances in transcription technologies have facilitated the transformation of spoken STEMM content into text suitable for computational analysis ^4^. These technologies have enabled the development of models that could capture linguistic subtleties inherent in STEMM-related discussions and extract valuable insights from expert dialogue. By transcribing and processing this informative podcast content, it has become feasible to augment LLMs with audio-linguistic data, providing a means of refining their understanding of domain-specific language, reasoning, and contextual interactions ^5^.

Here we present PodGPT (Fig. 1), a computational framework designed to leverage an extensive corpus of STEMM-based podcast content. Over 3, 700 hours of audio were transcribed into over 42 million text tokens, enabling the model to absorb and learn from diverse expert discussions across multiple scientific fields. By integrating spoken content, we aim to enhance the model’s understanding of conversational language and extend its application to more specialized contexts within STEMM disciplines. To further augment the model’s utility, we implemented a retrieval augmented generation (RAG) framework ^6,7^ utilizing a vector database built from articles in Creative Commons PubMed Central and *The New England Journal of Medicine* (NEJM). This database contained a heterogeneous and high-impact selection of medical and scientific literature, grounding PodGPT’s outputs in peer-reviewed knowledge. The RAG approach was optimized by applying hybrid search with binary quantization techniques to improve retrieval effectiveness and efficiency, followed by re-ranking to ensure accurate and context-aware responses. This computational framework allows PodGPT to retrieve and incorporate relevant scientific evidence into its generative process, ensuring linguistically coherent responses grounded by referenced sources.

**Figure 1:**
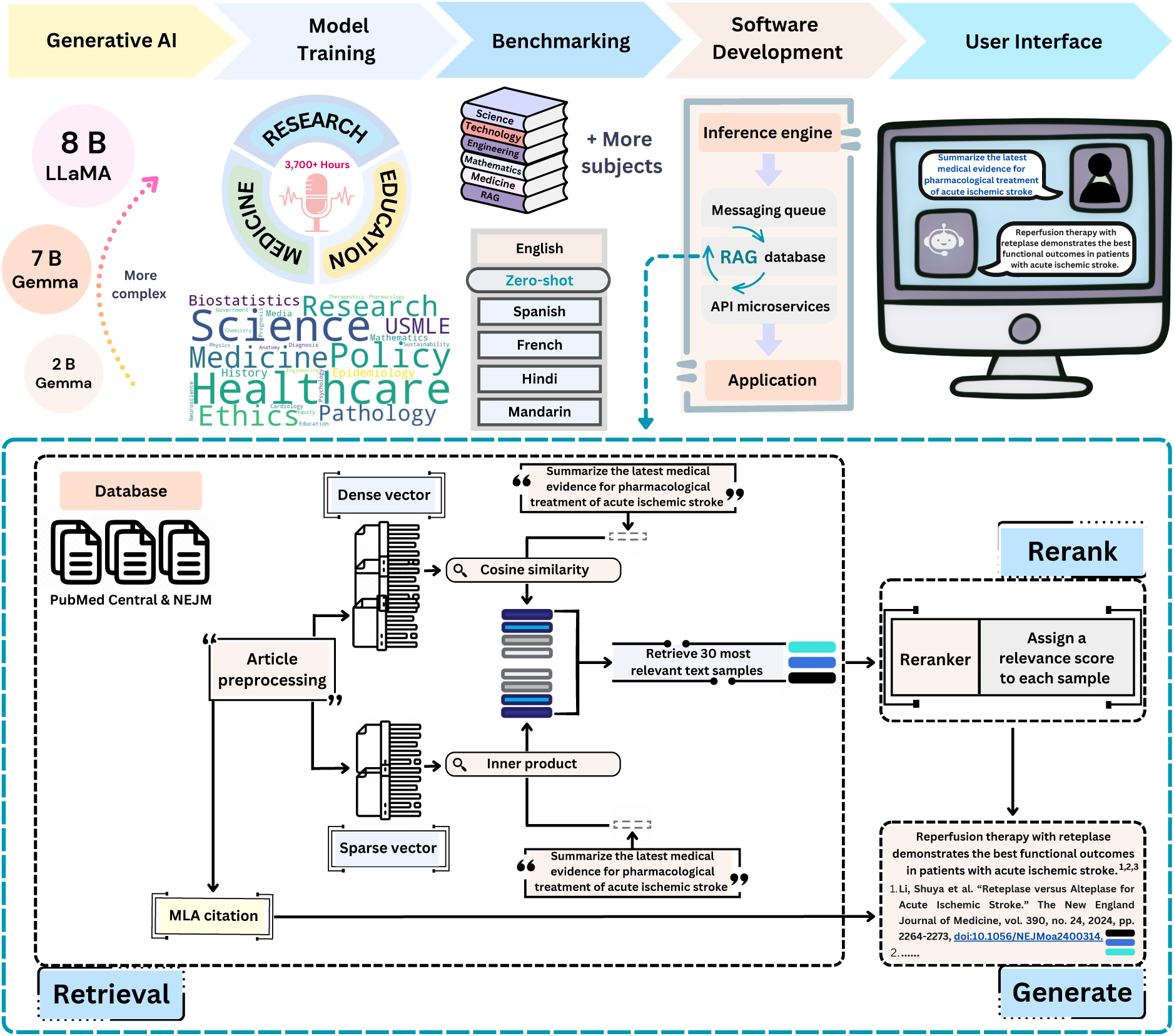
PodGPT framework. This figure illustrates the workflow and components involved in the development of PodGPT, an audio-augmented large language model tailored for research and education. The process began by leveraging publicly available generative AI auto-regressive language models across various scales. These models underwent continuous pre-training on a curated dataset of English CC-BY podcasts produced by scientific journals and clinical experts, as well as content from *The New England Journal of Medicine* (NEJM). The podcast corpus comprised over 3, 700 hours of audio, covering diverse topics in science, research, and medicine, visually summarized in the accompanying word cloud. Following pre-training, PodGPT was rigorously evaluated against leading English medical question-answering benchmarks, such as MedExpQA, MedMCQA, MedQA, and PubMedQA. It also underwent assessment on STEMM subsets within MMLU benchmarks, encompassing subjects like physics, biology, chemistry, computer science, engineering, mathematics, and medicine. Additionally, to evaluate its zero-shot transfer capability, multilingual STEMM benchmarks were included, covering widely spoken languages such as Spanish, French, Hindi, and Mandarin. The next phase involved developing the software infrastructure, which included an inference engine for model deployment, a messaging queue, database integration, retrieval augmented generation (RAG) implementation, API microservices, and a responsive human-machine interface. This highly performant and robust system enabled users with internet access to engage seamlessly with current research and educational material via an adaptive chatbot. The chatbot supported multi-turn conversations across various languages, empowering users to access and interact with STEMM knowledge in a dynamic and accessible manner.

The development of PodGPT positions it as a valuable tool for research and educational applications within STEMM, particularly in contexts requiring precise domain-specific knowledge. Its capacity to integrate complex interdisciplinary conversations into language models highlights the potential of LLMs to support knowledge dissemination and professional development across STEMM disciplines. By bridging the gap between static text-based corpora and dynamic audio content, PodGPT exemplifies a forward-thinking approach to refining the capabilities of language models for scientific inquiry.

## Methods

### Dataset description

We curated a diverse collection of podcasts across STEMM disciplines under the Creative Commons Attribution (CC-BY) license as well as content from *The New England Journal of Medicine* (NEJM). The CC-BY license supports content sharing, adaptation, and use for any purpose, provided appropriate credit is given. Various versions of the CC-BY license (*e*.*g*., 1.0, 2.0, 3.0, and 4.0) outline specific attribution requirements and compatibility updates with international copyright standards. The latest version, CC-BY 4.0, offers enhanced flexibility by allowing sharing and adaptation across jurisdictions, making it widely applicable in research and educational contexts, as exemplified by our use of these podcasts for training PodGPT.

Our curated corpus was comprised of podcasts produced by scientific journals, researchers, and clinicians, with topics spanning a multitude of different scientific fields. We filtered these podcasts based on the following criteria: (1) podcasts hosted by reputable scientific journals, such as NEJM; (2) podcasts produced by individuals with recognized scientific, medical, or research expertise, including medical doctors (M.D.) and doctors of philosophy (Ph.D.), that encompassed various STEMM fields. The full set of podcast episodes and associated metadata are detailed in Table 1.

**Table 1:** Podcasts used for model development. This table summarizes the STEMM podcasts licensed under Creative Commons Attribution (CC-BY) and content from *The New England Journal of Medicine* (NEJM) used for PodGPT’s continual pre-training. It provides key details, including podcast names, episode counts, total audio durations, average episode lengths with standard deviations, text token counts, and average tokens per episode with standard deviations. Podcasts from NEJM, *The Journal of Sustainable Development*, and *The Columbia University Journal of Global Health* offered a broad range of episodes with substantial audio durations and high token volumes, capturing in-depth discussions on critical topics in research and education. The podcast content, transcribed using OpenAI’s Whisper model, forms a diverse and comprehensive dataset that strengthens PodGPT’s knowledge base and comprehension across STEMM domains.

| Podcast | Episodes | Audio Time (min) | Mean Length Episode $\pm \sigma$ (min) | Number of Text Tokens | Mean Text Tokens per Episode $\pm \sigma$ |
| --- | --- | --- | --- | --- | --- |
| NEJM This Week | 457 | 13,300.17 | 29.10 ± 2.92 | 2,029,219 | 4440.30 ± 419.35 |
| NEJM Interviews | 654 | 9,223.44 | 14.10 ± 8.15 | 1,732,879 | 2649.66 ± 1522.35 |
| NEJM Core IM \| Internal Medicine Podcast | 170 | 5,285.72 | 31.09 ± 10.08 | 1,077,154 | 6336.20 ± 2093.18 |
| NEJM Curbside Consults | 74 | 1,977.46 | 26.72 ± 11.04 | 408,189 | 5516.07 ± 2522.46 |
| NEJM Clinical Conversations | 108 | 1,829.66 | 16.94 ± 4.13 | 320,968 | 2971.93 ± 774.89 |
| NEJM Leadership Conversations | 100 | 1,765.76 | 17.66 ± 4.38 | 306,490 | 3064.90 ± 759.09 |
| NEJM AI Grand Rounds | 24 | 1,459.11 | 60.80 ± 14.84 | 303,499 | 12645.79 ± 3107.35 |
| NEJM Intention to Treat | 40 | 997.22 | 24.93 ± 5.27 | 169,632 | 4240.80 ± 954.41 |
| NEJM Not Otherwise Specified | 20 | 836.03 | 41.80 ± 15.71 | 146,718 | 7335.90 ± 2880.36 |
| TWiV: This Week In Virology | 1,186 | 104,089.57 | 87.77 ± 30.30 | 20,188,268 | 17022.15 ± 5785.86 |
| TWiP: This Week In Parasitism | 245 | 21,129.03 | 86.24 ± 15.35 | 4,381,511 | 17883.72 ± 3814.75 |
| TWiM: This Week in Microbiology | 320 | 20,641.50 | 64.50 ± 10.36 | 3,667,519 | 11461.00 ± 2121.78 |
| TWiEVO: This Week In Evolution | 100 | 8,845.09 | 88.45 ± 10.99 | 1,756,480 | 17564.80 ± 2517.80 |
| IMMUNE | 93 | 6,918.84 | 74.40 ± 19.67 | 1,363,176 | 14657.81 ± 3888.47 |
| TWiN: This Week In Neuroscience | 53 | 3,581.13 | 67.57 ± 10.40 | 644,712 | 12164.38 ± 2078.49 |
| Matters Microbial | 62 | 3,418.33 | 55.13 ± 11.33 | 637,647 | 10284.63 ± 2357.42 |
| Infectious Disease Puscast | 65 | 2,298.71 | 35.36 ± 5.96 | 415,145 | 6386.85 ± 1126.15 |
| Urban Agriculture | 29 | 2,171.63 | 74.88 ± 17.58 | 443,859 | 15305.48 ± 4214.82 |
| On The Wards: On The Pods Medical Podcast for Doctors | 245 | 5,915.14 | 24.14 ± 8.00 | 1,175,307 | 4797.17 ± 1781.74 |
| Digital Campus Podcast | 64 | 3,169.44 | 49.52 ± 5.85 | 605,977 | 9322.72 ± 1528.99 |
| emDOCs.net Emergency Medicine (EM) Podcast | 112 | 1,576.72 | 14.08 ± 4.30 | 303,672 | 2711.35 ± 864.29 |
| Policy in Plain English Podcast | 73 | 1,089.20 | 14.92 ± 7.52 | 208,580 | 2857.26 ± 1509.80 |
| Open Minds ... from Creative Commons | 21 | 803.28 | 38.25 ± 12.79 | 145,624 | 6619.27 ± 2722.90 |
| What is Global Health? | 18 | 486.47 | 27.03 ± 10.02 | 87,653 | 4869.61 ± 2054.00 |
| Consilience Sustainability In Progress (SIP) Podcast | 9 | 403.95 | 44.88 ± 17.89 | 70,461 | 7829.00 ± 3535.75 |
| Research Pulse: Future Focussed Health Insights | 16 | 177.79 | 11.11 ± 2.29 | 33,672 | 2104.50 ± 476.85 |
| Our People: Central to Healthcare | 9 | 161.15 | 17.91 ± 7.42 | 31,545 | 3505.00 ± 1474.57 |

### Dataset processing

The pretraining corpus for PodGPT consisted of thousands of hours of STEMM podcasts, encompassing academic discussions, clinical case studies, and expert interviews. We transcribed these audio files using a state-of-the-art automatic speech recognition model, OpenAI Whisper ^8^. Built upon an encoder-decoder Transformer architecture, the Whisper model resampled the input audio to 16, 000 Hz and performed temporal chunking. Then, these chunks of audio data were represented by 80-channel log-magnitude Mel spectrograms with a 25-millisecond window and 10-millisecond stride. Before being processed by the Transformer modules, the input underwent a convolutional layer and was augmented with the sinusoidal position embeddings to incorporate positional information. Finally, the Transformer decoder module interpreted the hidden representation of the audio data and generated textual output through a language head ^9^. We utilized the latest Whisper series model, the Whisper large-v3, with 1, 550M parameters, to specify the spoken language for improved speech recognition. All the podcast transcripts were carefully and manually reviewed to maintain content quality.

### Model architecture

The Transformer model ^10^, renowned for its multi-head self-attention mechanism, has become the backbone of many state-of-the-art AI models. Unlike traditional methods, the self-attention mechanism captures long-range dependencies with efficient parallelization and scalability. Additionally, its deep feedforward neural networks enhance the model’s capacity to learn complex patterns in data. We leveraged this architecture to tailor PodGPT specifically for use in STEMM research and educational purposes. Built upon state-of-the-art general LLMs, such as Gemma ^11^ and LLaMA ^12^, PodGPT was pre-trained on a diverse and informative text corpus extracted from STEMM podcast data. By utilizing instruction-tuned variants of these models, we aimed to improve instruction-following capabilities and conversational structure. To evaluate the effectiveness of our framework, we applied models of varying scales, ranging from 2 to 8 billion parameters.

Gemma is a series of lightweight open models developed by Google DeepMind. These are text-to-text auto-regressive language models, which have pre-trained versions as well as instruction-tuned variants. These models were trained on the textual datasets on a context length of 8, 192 tokens from a wide variety of sources. The primary sources include web documents, codes, and mathematical content. Several recent advancements have been made to improve the performance and training efficiency of the Transformer model. These include multi-query attention ^13^, rotary positional embeddings (RoPE) ^14^, and GeGLU activations ^15^. We utilized the Gemma models 2B and 7B to validate our framework across different model sizes. LLaMA is a family of advanced general-purpose LLMs released by Meta Research. The LLaMA 3.1 8B was pretrained using a context length up to 128K, facilitating longer context understanding with RoPE. Additionally, it employs the standard decoder-only architecture with improved efficiency using grouped-query attention with 8 key-value heads ^16^.

Pre-training is a crucial step in the development of LLMs, during which the model learns from a vast body of text data in an auto-regressive manner. This phase generally leverages self-supervised learning, employing methods like masked language modeling, *e*.*g*., BERT ^17^, or autoregressive modeling, *e*.*g*., GPT ^18^. The self-supervised learning framework allows the model to gain a broad understanding of knowledge, thereby improving its performance in subsequent tasks. In this work, we utilized an auto-regressive objective to perform continual pretraining through an iterative gradient solver. The above-mentioned LLMs have been pre-trained on trillions of tokens. Thus, one cost-effective and efficient way to encode domain-specific knowledge is through continuous pre-training and evolving the pre-trained models with expertise corpora, instead of retraining them from scratch. The podcast transcripts were represented by a sequence of tokens, *i*.*e*., **x** = *{x*_1_*, x*_2_*, . . . x_N_ }*, where *x_i_* is a subword token and *N* denotes the length of the sequence. We fine-tuned publicly available models using our podcast data in an auto-regressive manner, optimizing the models by minimizing the negative log likelihood. The training objective is as follows,

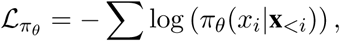

where *π_θ_*is the language model, parameterized by *θ*.

### Retrieval augmented generation framework

To ensure PodGPT is aligned with the latest advancements in STEMM research and education, we integrated a retrieval augmented generation (RAG) system into its computational framework. This system facilitates the continuous search and retrieval of up to date information from PubMed Central ^19^, the free archive of life science journal articles managed by the National Institutes of Health (NIH), United States. The NIH offered an FTP server for bulk downloading large sets of non-commercially licensed public articles, which we utilized to ground text generation in PodGPT. Additionally, the latest articles from NEJM since 2016 were also incorporated into our database, providing a wide range of content for medical research and education. The retrieved articles used to generate and ground the responses were meticulously cited in MLA format, ensuring proper attribution and ease of reference. This citation style facilitated clear acknowledgment of the original sources while maintaining the academic integrity of the outputs. By integrating these citations directly into the generated text, PodGPT ensured that users could trace back the evidence to peer-reviewed and credible scientific literature, further enhancing trustworthiness and usability for research and educational purposes.

To fully utilize the content, the first step was to preprocess article bodies into shorter text samples, determined by the hyper-parameters of the vectorization neural networks. Then, we encoded these text chunks using two embedding models. Specifically, we used two types of embeddings: a dense embedding, which is a compact vector that captures semantic similarity between different contexts, and a sparse vector, with the length of the vocabulary size, *e.g.*, 30K. In the sparse vector, each position corresponds to a subword token, enabling highly efficient keyword searches. We selected state-of-the-art open-source embedding models for both dense and sparse embeddings: the Salesforce Research SFR-Embedding-2 ^20^ and the opensearchneural-sparse-encoding-v1 model from the OpenSearch Project ^21^. To enhance efficiency, we implemented the 4-bit GPTQ quantization ^22^ on the Salesforce model. The similarity search was performed using cosine similarity for dense vectors and inner product for sparse vectors, as specified in these models.

Furthermore, to improve retrieval performance for the dense vector, we employed a two-stage retrieve-rerank approach. In this method, we initially conducted binary quantization for top-*K* retrieval, returning the most relevant top-*N* documents from our 4-bit quantized model. Then, we re-ranked the results based on similarity scores using a reranker. The samples that had the shortest distance were re-ranked and passed into the reranker model, which assigned relevance scores to each document. We selected the BAAI general embedding model, bge-reranker-large ^23^, a publicly available top-performing re-ranking model. Reranking allows a hybrid search that combines specialized embedding models for document retrieval with the generation of re-ranked relevance scores for the model’s use.

The database, vector search, and indexing were all managed by a vector database system pgvector ^24^, a PostgreSQL extension that implements vector distance searches and the hierarchical navigable small worlds (HNSW) algorithm with standard SQL tools, enabling efficient pipeline development with familiar APIs.

### Experimental settings

To analyze the capabilities of PodGPT, we employed various model sizes and conducted extensive experiments across STEMM fields in a multilingual context. In current literature, benchmarks for multiple-choice question-answering (QA) were commonly utilized to evaluate the performance of large language models. Thus, in this work, we utilized the multilingual multiple-choice QA benchmarks to evaluate the model’s performance. In addition, we conducted experiments and documented the performance of all the models that were used in this study on multilingual STEMM benchmarks. This potentially advances the field with an open-source and unified multilingual benchmarking library covering training, inference, answer extraction, performance evaluation, real-world model deployment, as well as a pipeline of RAG for evidence-based medicine (EBM). Furthermore, to guarantee scientific reproducible research, we implemented all our experiments with a set of unified hyper-parameters. Thus, our work was out of the box without any specific hyper-parameter tuning and further optimization for different models.

### Evaluation benchmarks

To evaluate the performance of PodGPT, we utilized a comprehensive set of STEMM benchmarks from the most spoken languages in the world, including English, Mandarin, French, Spanish, and Hindi. For intra-language experiments, *i*.*e*., English, we performed performance evaluations on datasets where the language aligned with the podcast content. Furthermore, for cross-language experiments, the model was evaluated on benchmarks in different languages compared to the podcasts. This evaluation was crucial for validating the effectiveness of the zero-shot multilingual transfer capability of medical LLMs. Our multilingual benchmarking approach not only demonstrates that our model is accurate and effective across varied linguistic contexts, but that it represents a technical achievement with the power to democratize global access to science, research, and educational knowledge. The detailed descriptions of multilingual benchmarks are as follows.

#### STEMM benchmarks in English

The benchmarks for STEMM natural language understanding in English have advanced significantly over the past decade. In this study, we selected well-known publicly accessible benchmarks including MedQA ^25^, PubMedQA ^26^, MedMCQA ^27^, MedExpQA ^28^, and MMLU STEMM datasets, including subsets from physics (astronomy, college physics, conceptual physics, and high school physics), biology (college biology and high school biology), chemistry (college chemistry and high school chemistry), computer science (college computer science, computer security, high school computer science, and machine learning), engineering (electrical engineering), mathematics (abstract algebra, college mathematics, elementary mathematics, high school mathematics, and high school statistics), and medicine (anatomy, clinical knowledge, college medicine, medical genetics, professional medicine) ^29^. Additionally, we incorporated the college biology question set into the MMLU medicine subset to ensure a fair comparison, as it was widely used to evaluate large medical language models ^30^.

#### STEMM benchmarks in Mandarin

The benchmarking of medical and clinical knowledge in Mandarin has become increasingly popular in NLP research. A range of databases have been successively proposed to assess the performance of Mandarin language models on medical data. In this study, we adopted the popular MedQA-MCMLE ^25^ and CMMU STEMM topics ^30,31^. The CMMLU STEMM benchmarks include the subsets from physics (astronomy, conceptual physics, and high school physics), biology (high school biology), chemistry (high school chemistry), computer science (computer science, machine learning, and computer security), mathematics (college actuarial science, college mathematics, elementary mathematics, high school mathematics, and college medical statistics), engineering (college engineering hydrology and electrical engineering), and medicine (anatomy, clinical knowledge, college medicine, genetics, nutrition, traditional Chinese medicine, and virology) ^31^.

#### STEMM benchmarks in Spanish

The Spanish STEMM testbed has encouraged the NLP community to develop new approaches for understanding and reasoning about science, research, and educational knowledge in Spanish. Therefore, we utilized the HEAD-QA benchmark, a multiple-choice healthcare dataset obtained from examinations in the Spanish healthcare system ^32^. Additionally, we also employed the MedExpQA Spanish subset ^28^ and Spanish MMLU STEMM topics ^30^.

#### STEMM benchmarks in Frenc

We primarily selected the popular FrenchMedMCQA dataset, which consists of 3, 105 questions taken from the French pharmacy diploma examinations ^33^. Following Wang et al., we only performed performance evaluations on questions with a single answer ^30^. As a result, the total number of questions in the testing set was 321. Furthermore, the MedExpQA French subset ^28^ and French MMLU STEMM topics ^30^ were also included in this work.

#### STEMM benchmarks in Hindi

To encode STEMM content in Hindi, we included the Hindi MMLU STEMM topics ^30^ in our benchmarking. By evaluating our model’s ability to understand science, research, and medical terminology in Hindi, we were able to incorporate one of the most widely spoken languages in the world.

### Implementation details

We transcribed podcast data using the OpenAI Whisper large-v3 model for an automatic speech recognition task. The chunk length was set to 30 seconds with a 5-second stride on both sides to improve the continuity and coherence of the transcriptions. The batch size was 96, and 384 tokens were generated per chunk to parallelly process audio chunks.

We encoded STEMM knowledge across various model sizes, from 2B to 8B. To do so, we imple-mented publicly available language models, which include the 2B and 7B versions of the Gemma series and the instruction-tuned variant of the LLaMA 3.1 8B. During model training, we utilized Brain float 16 data type with the AdamW optimizer to prevent overflow issues ^34^, and the context length was set to 2, 048 ^30^.

We trained all models for 5 epochs with an initial learning rate of 5 *×* 10^−6^ with a 0.03 warm-up ratio and a cosine schedule. The weight decay rate was 0.01, and the gradient was accumulated during each training step. All the models were optimized based on the unified hyper-parameter settings without specific tuning for superior performance. To deploy a highly performant and efficient RAG pipeline, we selected the top*K* = 10, 000 document samples using the binary quantized embedding model. Next, we retrieved the text samples from the top-*N* = 15 dense embeddings and the top-*N* = 15 sparse embeddings by similarity score. Finally, we used only the documents that had reranking scores greater than 1.0.

### Software and database infrastructure

We created a custom graphical user interface (GUI) and platform infrastructure to allow users to interact with PodGPT, providing public access to our model. Our goal was to deliver our model with a user-friendly and responsive conversational interface. We utilized ReactJS and NextJS for the front end. ReactJS furnishes a collection of APIs and libraries to construct reusable web components, while NextJS provides scaffolding for ReactJS applications, encompassing an HTTP server, server-side rendering, and a “back end for a front end” design pattern. For hardware, we utilized a custom-built stack to deploy our LLMs at scale using entirely self-hosted and open-source tools without relying on software as a service (SaaS) or proprietary software. We employed a microservice architecture using Kubernetes as a container orchestration tool. Kubernetes manages clusters of nodes hosting microservices wrapped inside Docker containers. It facilitates the creation of highly available distributed systems that automatically scale to meet needs and ensure secure inter-cluster communication, IP address allocation, load balancing, and reverse proxy services. For LLM deployment, we employed the vLLM library ^35^, which offers a fast and portable inference server that deploys transformers and batches inference tasks efficiently while providing a convenient API. It requires a minimum of CUDA Toolkit 11.0 and a 7.5 compute-capable NVIDIA GPU, supporting splitting model weights across several devices.

Authentication and user management were crucial components of our architecture. In order to distribute resources equitably among potential researchers, we implemented OAuth 2.0 compliant authorization and user management in addition to a per-token rate limiting system based on user scopes and total system load. We equipped PodGPT with standard chatbot features such as multiturn conversations and the ability to open new conversations. Furthermore, PodGPT utilized Apache Cassandra, a distributed NoSQL database designed for high availability and query optimization. The backend API router, which was built with Flask, stores new chats and conversations in Cassandra and sends text inference requests to a queue. For queuing and message processing, we utilized RabbitMQ and Redis, which are a message broker and key-value databases, respectively.

### Data and model availability

A multilingual LLMs benchmarking library along with the source codes are made available at github.com/vkola-lab/PodGPT. Our models are available at huggingface.co/vkola-lab.

## Results

We conducted comprehensive experiments to assess PodGPT’s performance on various multilingual STEMM QA benchmark datasets. Our results broadly demonstrate that incorporating STEMM audio podcast data enhances our model’s ability to understand and generate precise and comprehensive information. In addition, our enhanced models outperformed their respective baselines across a wide range of scales in both in-domain benchmarks and zero-shot domain generalization across multilingual benchmarks.

### Performance on in-domain benchmarks

The evaluation of PodGPT across a wide spectrum of disciplines in MMLU benchmarks demonstrated enhanced model efficacy following pre-training with STEMM podcast data (Table 2). Across all MMLU STEMM benchmarks, PodGPT achieved performance gains of 9.73 percentage points for the LLaMA 8B model and 3.49 percentage points for the Gemma 7B model. Specifically, on the MMLU physics benchmark subset, PodGPT outperformed the baseline by 12.36 percentage points. In MMLU biology benchmarks, there were 8.62 percentage points improvement for the LLaMA 8B model and 3.11 for the Gemma 7B model. Additionally, the Gemma 2B model showed enhancements of 3.61 percentage points on the MMLU computer science datasets. Evaluation of the engineering subset revealed that PodGPT achieved improvements of 10.34 and 3.80 percentage points for the LLaMA 8B and Gemma 7B models, respectively. Moreover, on the MMLU mathematics benchmark, PodGPT consistently outperformed the baseline, with the Gemma 7B model improving by 4.26 percentage points and the LLaMA 8B model improving by 4.25 percentage points. Lastly, on the medical datasets, PodGPT exhibited improvements of 10.29 percentage points for the LLaMA 8B model and 5.26 for the Gemma 7B model. Overall, PodGPT demonstrated cumulative enhancements of 3.51 percentage points over standard open-source benchmarks and 3.13 percentage points across in-domain MMLU benchmarks. These results underscored the potential of leveraging open-source podcast data to significantly boost model performance and applicability across specialized STEMM domains, paving the way for more effective and versatile AI systems for research and education.

**Table 2:** Performance of PodGPT on English benchmarks. All models were fine-tuned using English podcast data and evaluated on STEMM subsets within MMLU benchmarks, which include physics, biology, chemistry, computer science, engineering, mathematics, and medicine. The performance of baseline models was compared against our PodGPT model (denoted as *Ours*). Bold numbers highlight the best-performing model in each category, showcasing PodGPT’s ability to achieve superior results across various STEMM domains.

| Language | MMLU Benchmark | Model |  |  |  |  |  |
| --- | --- | --- | --- | --- | --- | --- | --- |
|  |  | Gemma 2B |  | Gemma 7B |  | LLaMA 8B |  |
|  |  | Baseline | *Ours* | Baseline | *Ours* | Baseline | *Ours* |
| English | Physics | 29.85 | **30.74 (0.12)** | 42.38 | **47.64 (0.64)** | 49.83 | **58.46 (0.36)** |
|  | Biology | 44.82 | **46.58 (0.45)** | 63.32 | **66.43 (0.58)** | 69.06 | **78.18 (0.22)** |
|  | Chemistry | 30.04 | **30.96 (0.62)** | 43.42 | **44.14 (1.31)** | 48.08 | **50.82 (1.08)** |
|  | Computer Science | 40.59 | **44.20 (0.11)** | 53.58 | **54.62 (0.29)** | 53.10 | **57.22 (0.33)** |
|  | Engineering | 39.31 | **42.07 (0.49)** | 44.14 | **47.94 (0.60)** | 48.97 | **59.31 (0.49)** |
|  | Mathematics | 25.69 | **26.44 (0.57)** | 34.97 | **39.23 (0.16)** | **50.41** | 43.13 (0.62) |
|  | Medicine | 40.62 | **41.72 (0.15)** | 55.22 | **59.50 (0.14)** | 66.11 | **74.05 (0.16)** |
|  | Average | 34.92 | **36.36 (0.11)** | 47.66 | **51.15 (0.27)** | 55.89 | **59.84 (0.13)** |

As shown in Table 3, we further evaluated PodGPT across various English medical QA benchmarks after pre-training with English medical podcast data with the RAG pipeline. This framework consistently surpassed baseline models, achieving an improvement of up to 13.00 percentage points, highlighting the effectiveness of grounding language models with the latest scientific evidence. On the MedExpQA bench-mark, our framework demonstrated an improvement of 12.20 percentage points for the Gemma 7B model. The other models also exhibited strong performance enhancements of 4.80 and 2.80 percentage points for the Gemma 2B and LLaMA 8B models, respectively. Additionally, PodGPT with RAG excelled on the MedQA benchmark, achieving an average increase of up to 4.04 percentage points. On the MMLU medicine bench-mark, promising improvements were observed, with gains of 5.97 and 3.54 percentage points for the Gemma 7B and LLaMA 8B models, respectively. Finally, on the MedMCQA benchmark, the Gemma 7B model with RAG surpassed the baseline by 2.90 percentage points. Overall, PodGPT with RAG demonstrated a cumulative enhancement of 3.81 percentage points across in-domain medical benchmarks, emphasizing its effectiveness in advancing medical education and research through the incorporation of evidence-based medicine with RAG.

**Table 3:** Performance of PodGPT with RAG on English benchmarks. All models were fine-tuned with English STEMM podcast data and evaluated on various medical QA benchmarks, including MedExpQA, MedMCQA, MedQA, PubMedQA, and the MMLU Medicine subset. For each model, baseline performance was compared against PodGPT (indicated as *Ours*). Additionally, the performance of baseline models integrated with RAG (denoted as *Baseline+RAG*) and PodGPT with RAG (denoted as *Ours+RAG*) was evaluated. The results demonstrated PodGPT’s superior performance, highlighting the effectiveness of incorporating podcast data into the training process. Bold numbers indicate the best-performing model in each category.

| Benchmark Datasets |  |  | MedExpQA | MedMCQA | MedQA | PubMedQA | MMLU Medicine | Average |
| --- | --- | --- | --- | --- | --- | --- | --- | --- |
| Model | Gemma 2B | Baseline | 19.20 | **34.71** | 29.54 | 46.80 | 40.62 | 34.17 |
|  |  | *Ours* | **21.20 (0.69)** | 34.62 (0.02) | **32.91 (0.15)** | **54.25 (0.54)** | **41.72 (0.15)** | **36.94** |
|  |  | *Baseline+RAG* | 23.20 | 35.91 | 32.60 | 49.00 | 41.47 | 36.44 |
|  |  | *Ours+RAG* | **28.00 (0.00)** | **35.96 (0.07)** | **34.43 (0.12)** | **51.95 (0.78)** | **42.12 (0.13)** | **38.49** |
|  | Gemma 7B | Baseline | 34.40 | 40.69 | 37.78 | **61.80** | 55.22 | 45.98 |
|  |  | *Ours* | **42.00 (0.89)** | **44.64 (0.09)** | **44.14 (0.21)** | 57.35 (1.37) | **59.50 (0.14)** | **49.53** |
|  |  | *Baseline+RAG* | 35.20 | 40.64 | 39.28 | **61.40** | 54.14 | 46.13 |
|  |  | *Ours+RAG* | **47.40 (1.18)** | **43.54 (0.07)** | **43.32 (0.25)** | 55.70 (1.88) | **60.11 (0.39)** | **50.01** |
|  | LLaMA 8B | Baseline | 55.20 | 51.28 | 53.50 | 72.00 | 66.11 | 59.62 |
|  |  | *Ours* | **62.20 (0.35)** | **55.90 (0.10)** | **59.80 (0.25)** | **73.75 (0.22)** | **74.05 (0.16)** | **65.14** |
|  |  | *Baseline+RAG* | 54.40 | 53.93 | 54.75 | 72.40 | 68.13 | 60.72 |
|  |  | *Ours+RAG* | **57.20 (0.40)** | **54.98 (0.16)** | **57.07 (0.28)** | **72.50 (0.22)** | **71.67 (0.21)** | **62.68** |

### Zero-shot cross-lingual performance

In Table 4, we presented results on PodGPT’s zero-shot crosslingual performance using multilingual benchmarks, encompassing diverse STEMM subjects such as physics, biology, chemistry, computer science, engineering, mathematics, and medicine. PodGPT demonstrated performance gains across all benchmarks, achieving improvements of up to 7.06 percentage points. The LLaMA 8B model achieved an improvement of 13.60 percentage points on the MedQA-MCMLE benchmark, show-casing its strong cross-lingual capabilities. Additionally, it delivered superior performance on the CMMLU benchmarks, achieving average improvements of up to 6.80 percentage points. The LLaMA 8B model demonstrated outstanding performance gains of 19.99, 11.53, 10.34, and 10.32 percentage points on CMMLU benchmarks focusing on chemistry, mathematics, physics, and computer science, respectively. Furthermore, the Gemma 7B model achieved increases of 7.57 and 7.14 percentage points on the CMMLU chemistry and computer science benchmarks, separately, highlighting the model’s robustness across diverse STEMM disciplines.

**Table 4:**
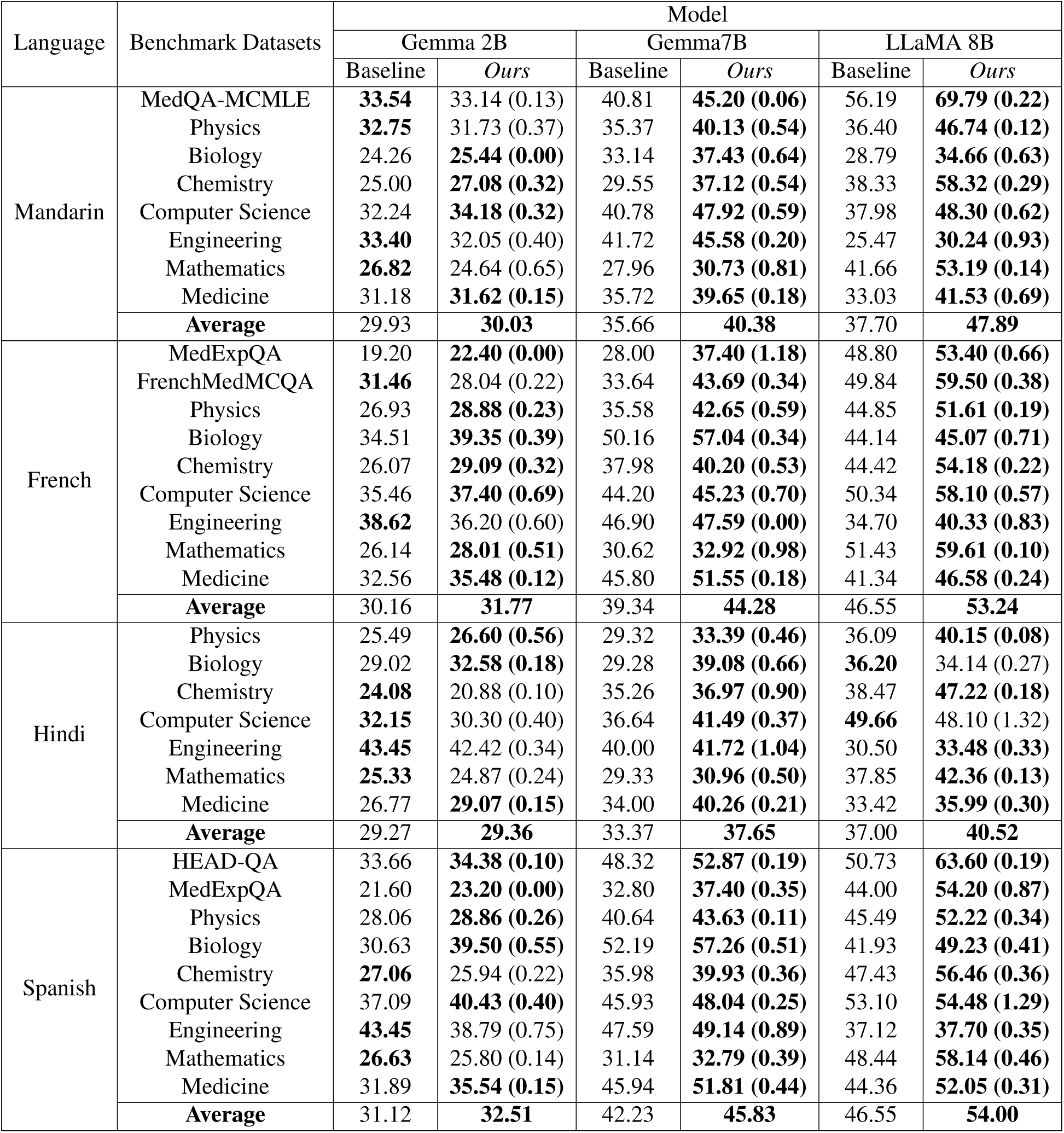
Zero-shot performance of PodGPT on non-English benchmarks. All models were finetuned using English podcast data and evaluated on various multilingual STEMM QA benchmarks in languages such as Mandarin, French, Hindi, and Spanish. These benchmarks included MedQA-MCMLE, FrenchMedMCQA, MedExpQA, HEAD-QA, and STEMM subsets within MMLU and CMMLU, covering physics, biology, chemistry, computer science, engineering, mathematics, and medicine. The performance of baseline models was compared to that of our model, PodGPT (denoted as *Ours*), to highlight the impact of integrating podcast data into the training process. Bold numbers indicate the superior performance in each category.

On the MedExpQA benchmark, PodGPT achieved performance gains of 9.40 percentage points for the Gemma 7B model and 4.60 percentage points for the LLaMA 8B model. It also outperformed the baseline model on the MedMCQA benchmark, with improvements of 10.05 percentage points for Gemma 7B and 9.66 for LLaMA 8B. In addition, the LLaMA 8B model showcased an average improvement of 8.41 percentage points across French STEMM benchmarks, while the Gemma 7B model achieved an average increase of 4.01 percentage points. Specifically, LLaMA 8B demonstrated an improvement of 9.76 percentage points on the chemistry subset, 8.18 on the mathematics subset, 7.84 on the computer science subset, and 5.63 on the engineering subset. On the French MMLU physics and biology subsets, PodGPT showed improvement, with gains of 7.07 and 6.88 percentage points for the Gemma 7B model. Additionally, PodGPT consistently surpassed the baseline models, achieving improvements of 5.75 percentage points with the Gemma 7B model and 5.24 percentage points with the LLaMA 8B model.

On the Hindi MMLU STEMM benchmarks, PodGPT achieved a performance gain, with an increase of 8.86 percentage points for the LLaMA 8B model. Additionally, the Gemma 7B model showed improvements of 9.80 on the biology benchmark, while the LLaMA 8B model beat the baseline by 8.75 percentage points for the chemistry benchmark. Additionally, on the Hindi MMLU medicine benchmark, PodGPT demonstrated an improvement of 6.26 percentage points for the Gemma 7B model. Lastly, the performance on the Spanish STEMM benchmarks was equally promising, with improvements of 12.87 percentage points and 10.20 percentage points on the HEAD-QA and MedExpQA benchmarks, respectively, for the LLaMA 8B model. Across the Spanish MMLU STEMM benchmarks, PodGPT achieved enhancements of up to 9.70 percentage points. Overall, PodGPT demonstrated its superiority by enhancing its zero-shot multilingual transfer capability in Spanish, achieving an average improvement of up to 7.45 percentage points. In summary, PodGPT showcased an impressive average improvement of 4.06 percentage points across multilingual STEMM benchmarks in its zero-shot transfer capabilities. These results underscored PodGPT’s effectiveness in generalizing across diverse linguistic contexts, further highlighting its potential for advancing multilingual applications in research and education.

### Evaluation of subject-specific queries

Our PodGPT model, integrated with the RAG framework, successfully generated up to date, relevant, and citation-supported responses to specialized STEMM queries, as demonstrated in Fig. 2. This integration empowered PodGPT to leverage a vector database of scientific literature, enabling the delivery of evidence-based answers underpinned by peer-reviewed references. Each response contained a relevance score, ensuring alignment between retrieved references and the query, and prioritizing highly pertinent sources while minimizing unrelated content. This approach significantly bolstered the reliability and precision of the generated outputs, as evidenced by consistently high relevance scores across diverse STEMM examples. In the examples shown in Fig. 2, we showed the effectiveness of PodGPT in addressing queries spanning endocrinology, infectious diseases, cardiovascular health, neuroscience, and planetary health.

**Figure 2:**
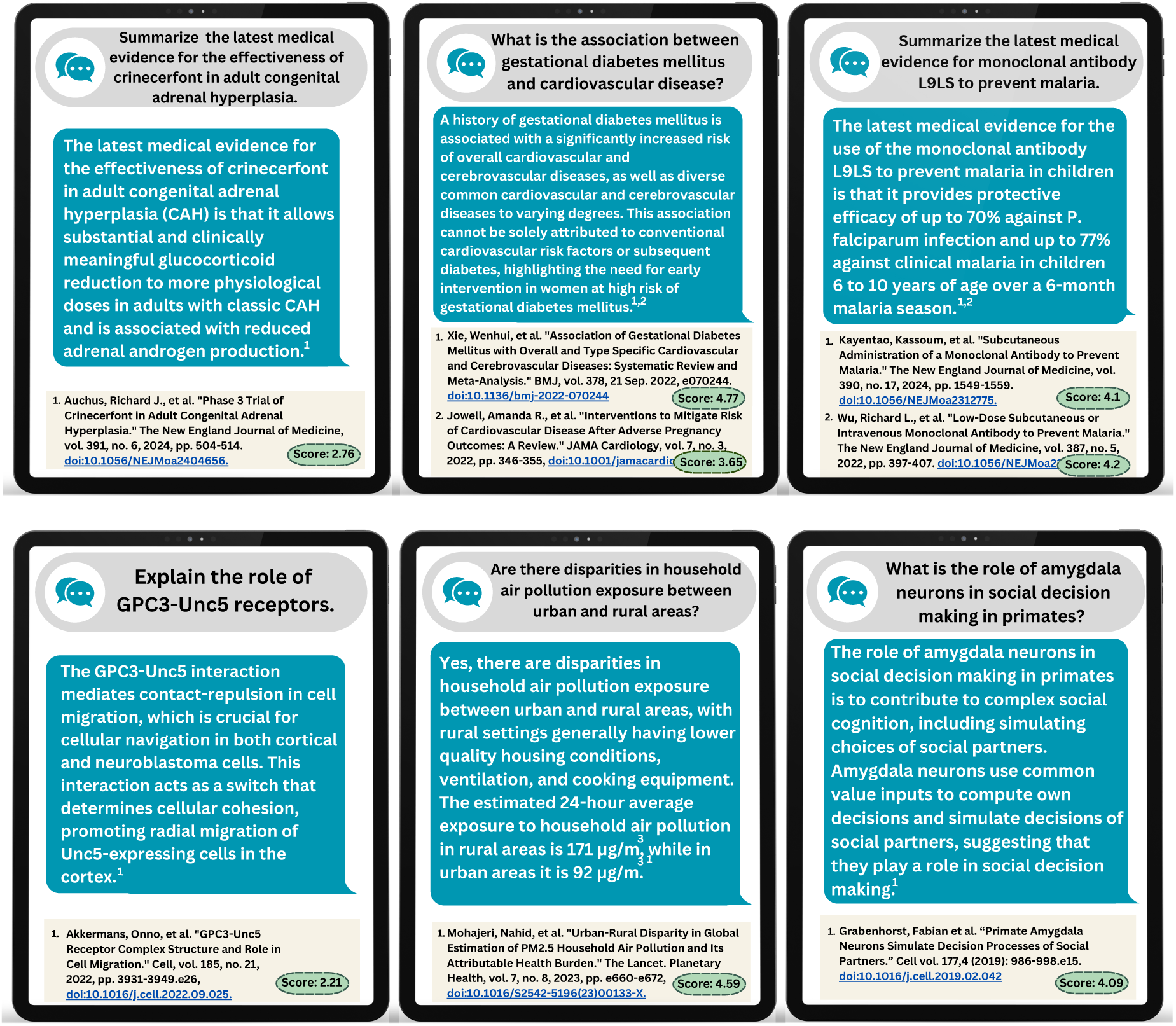
PodGPT responses on STEMM queries. The evaluation of PodGPT’s outputs using the retrieval augmented generation (RAG) framework highlighted its capacity to provide accurate, contextually relevant responses grounded by scientific references. The references were retrieved from a vector database and scored for relevance to the query using the *bge-reranker-large model*. The relevance score quantified the alignment between the query and retrieved references. A higher score indicated a stronger contextual match, with the inclusion of references determined by a tunable score threshold. This hyperparameter allowed for customization to optimize performance based on specific application needs. The examples provided illustrated PodGPT’s adeptness at generating precise and contextually grounded outputs across diverse STEMM topics, showcasing its reliability in integrating evidence-based information into its responses. This approach underscored the potential of PodGPT to enhance research and education by offering high-quality and citationsupported answers.

## Discussion

We present PodGPT, a large language model that leverages the rich and diverse linguistic content of STEMM podcasts, capturing a wide array of domain-specific terminologies and conversational contexts. Extensive pre-training on podcast data has endowed PodGPT with the capability to generate topically relevant and scientifically up to date responses to highly specialized STEMM-related queries across different languages. When benchmarked against existing datasets such as MedQA, PubMedQA, MedMCQA, and various MMLU STEMM categories, PodGPT demonstrated superior performance, particularly in areas requiring detailed medical knowledge and contextual understanding. We leveraged a RAG framework with a vector database constructed from journal articles, enabling real-time access to emerging scientific literature. These results not only highlight its potential as a valuable tool for research and education, but also to democratize science globally through its accessibility and multilingual capabilities.

Our results demonstrate that integrating audio-transcribed data into language model training improves the accuracy and relevance of information generated, particularly in STEMM contexts. Compared to baseline models such as Google Gemma and Meta LLaMA, PodGPT consistently achieved higher performance across multiple benchmarks, including in-domain STEMM tasks and zero-shot multilingual evaluations. By leveraging transcribed audio, PodGPT effectively captures conversational dynamics, domain-specific terminologies, and interdisciplinary dialogue patterns that are often absent in text-only training data. This integration enables more precise language processing and a broader understanding of complex topics. The implications of this work extend beyond benchmark performance. First, PodGPT underscores the importance of integrating diverse modalities, such as audio-transcribed text, into language model training to address linguistic subtleties and interdisciplinary dialogue patterns that static text corpora alone cannot encapsulate. This inclusion strengthens the model’s robustness, enabling not only superior performance on standard benchmarks but also enhanced generalization to multilingual and multidisciplinary applications. Second, the integration of a RAG framework equips PodGPT with real-time access to evolving scientific literature, providing researchers, educators, and practitioners with evidence-grounded insights that remain current and actionable. This capability bridges the gap between traditional language model outputs and dynamic, evidence-based decision-making, particularly in fast-evolving STEMM fields. Finally, PodGPT’s demonstrated success across multilingual STEMM benchmarks positions it as a transformative tool for democratizing access to education and research globally. By breaking down language and geographic barriers, PodGPT has the potential to promote equitable access to scientific knowledge, enabling underserved and linguistically diverse communities to engage with cutting-edge research. As the global demand for interdisciplinary collaboration and education grows, PodGPT serves as an example of how language models can evolve to meet these challenges by integrating innovative data modalities and frameworks.

Our study has a few limitations. First, we were limited to using STEMM podcast content that was openly accessible or publicly available. As such, there is a vast collection of available but unharnessed data, such as textbooks and even video tutorials, that could be leveraged if licensing guidelines allowed. Our model, trained exclusively on English podcasts, demonstrated strong performance on established benchmarks and improved zero-shot capabilities on multilingual evaluation tasks. Future efforts will focus on acquiring diverse and legally accessible podcast data in multiple languages as well as more peer-reviewed journal content to enrich training and enhance multilingual model performance. Future work on PodGPT should also include a comprehensive ethical evaluation to ensure the model consistently adheres to high scientific and research standards in a wide range of settings. Also, we observed that pre-training using podcast data did not improve performance on a few benchmarks. This finding can be attributed to the nature and structure of podcasts, which contrasts with the demands of these benchmarks.

The findings from this study indicate that PodGPT represents an important advancement in tailoring large language models for research and education. Its true potential, however, lies in democratizing access to scientific information globally. Its ability to process domain-specific queries, generate responses using the most up to date information available, including in-chat citations, and operate across multiple languages makes PodGPT a valuable tool. Nonetheless, deploying these advanced models must be accompanied by rigorous attention to data integrity and user privacy considerations. By continuing to advance the intersection of AI and science, we can ultimately improve the global accessibility of STEMM research and education, ensuring that such technologies benefit a broader range of people. PodGPT highlights the value of integrating podcast data to enhance language models, which can be extended to applications beyond just science and research by incorporating diverse audio podcasts.

## Data Availability

All data produced are available online at https://github.com/vkola-lab/PodGPT.

## Acknowledgments

This project was supported by grants from the Karen Toffler Charitable Trust (V.B.K.), the National Institute on Aging’s Artificial Intelligence and Technology Collaboratories (P30-AG073104 and P30-AG073105, V.B.K.), the American Heart Association (20SFRN35460031, V.B.K. & R.A.), Gates Ventures (R.A. & V.B.K.), and the National Institutes of Health (R01-HL159620 [V.B.K.], R01-AG083735 [R.A. & V.B.K.], R01-AG062109 [R.A. & V.B.K.], and U19-AG068753 [R.A.]).

## Author contributions

S.J., S.B., and E.S. contributed equally to this work. S.J., S.B., L.A.C., P.F., V.H.J., M.V.L., and D.V. curated and processed the data. S.J. and S.B. performed model training. E.S. developed the RAG framework. S.J., S.B., L.A.C., P.F., V.H.J., M.V.L., E.S., D.V. and W.M.W. generated the results. R.A. provided clinical and scientific context. V.B.K. drafted the initial manuscript. All authors reviewed, edited, and approved the manuscript. V.B.K. conceived, designed, and directed the study.

## Ethics declarations

V.B.K. is a co-founder and equity holder of deepPath Inc. and CogniScreen, Inc. He also serves on the scientific advisory board of Altoida Inc. R.A. is a scientific advisor to Signant Health and NovoNordisk. The remaining authors declare no competing interests.

